# Reductions and pronounced regional differences in morphine distribution in the United States

**DOI:** 10.1101/2022.05.16.22275134

**Authors:** Megan E. Dowd, E. Jessica Tang, Kurlya T. Yan, Kenneth L. McCall, Brian J. Piper

**Affiliations:** Geisinger Commonwealth School of Medicine, Scranton, PA; University of New England, Portland, ME; Center for Pharmacy Innovation & Outcomes, Forty Fort, PA

## Abstract

**Background:** Morphine is one of the oldest, most commonly prescribed, and widely used opioids in the United States (US). Morphine’s potent analgesic properties have also been associated with the increase in misuse, addiction and opioid-related deaths in the US since the 1990s. Despite federal regulations, population-adjusted prescription opioid distribution varies markedly between states. The objective of this study was to describe the temporal pattern of morphine distribution nationally and between states.

**Methods:** Drug weight and population data were obtained from Report 5 of the US Drug Enforcement Administration’s Automation of Reports and Consolidated Orders System (ARCOS) to characterize patterns in the distribution of morphine from 2012 to 2020.

Morphine distribution amounts were separated by state and business type and corrected for population. States outside a 95% confidence interval relative to the national average were considered statistically significant.

**Results:** Pharmacies and hospitals distributed 24,200 kilograms of morphine in 2012. Tennessee (180.2 mg/person) was 4.7-fold higher than Texas (39.4 mg/person).

National distribution decreased 56.4% to 10,723 kilograms in 2020. Tennessee (56.4 mg/person) was 3.8-fold higher than the District of Columbia (15.0 mg/person). The decline in Illinois (−40.9%) was significantly less than the national average (−56.8%) while that of Oregon (−71.1%) and Arizona (−70.4%) were significantly higher. Hospital decrease (−72.7%) from 2012-2020 was larger than that of pharmacies (−56.12%).

**Conclusions:** The national 56% decline in the distribution of morphine in the last decade may be attributable to prioritization of the opioid crisis as a public concern, including subsequent growth of opioid misuse and treatment programs and decreased production quotas for opioids, including morphine. This decline also coincides with the national shortage of parenteral opioids resulting in greater prescriptions of alternative opioids such as nalbuphine and buprenorphine. Further research is necessary to understand the persistent four-fold regional difference between states.

## Introduction

Morphine, a potent analgesic commonly prescribed for around-the-clock management of moderate to severe pain, is one of the most commonly abused opioids in the US (1) and, by weight, is the number two prescription opioid worldwide (2). The wide variety of formulations and strengths enables broad use for acute and chronic pain not only for adults but also for a wide variety of pediatric (age > 2) indications (3). In contrast, many opioids are only US FDA approved for adults. It is commonly prescribed for many conditions that cause acute and/or chronic pain, most notably in the palliative/end-of-life and postoperative care setting, as well as in the emergency department and cath lab (1). Adverse effects include increased risk of infections, including COVID-19 and COVID-19-related complications, increased risk of falls and fractures among elderly patients, neonatal opioid withdrawal syndrome, and death (1, 4). The varied routes of administration (oral, intravenous, epidural, intrathecal, sublingual, nasal) create greater opportunity for misuse. Treating opioid misuse is estimated to cost $78.5 billion each year, comparable to medical costs for asthma and diabetes (5). Opioid prescriptions increased by 64% between 2004-2011 but, after peaking in 2012, have since declined (6).

Despite an overall national decrease in morphine prescriptions, there was marked variation between states concerning the percent change in population-adjusted opioid distribution since the peak of prescription opioid distribution, including morphine usage, in 2011 (7). Substantial regional disparities have been identified for other opioids and may warrant additional attention including for improvements in opioid stewardship (7, 8, 9, 10, 11). Therefore, our objectives in this study were to: 1) describe national patterns in morphine distribution from 2010-2020 and 2) compare these at a state level.

## Methods

### Procedures

All data was collected through the Automation of Reports and Consolidated Orders System (ARCOS). ARCOS is an automated, comprehensive drug reporting system that allows the US Drug Enforcement Administration (DEA) to monitor the flow of controlled substances from the point of manufacture through commercial distribution channels to point of sale or distribution at the dispensing/retail level—hospitals, retail pharmacies, practitioners, mid-level practitioners, and teaching institutions (12). ARCOS tracks controlled substances transactions and monitors the distribution of controlled substances by weight (grams). The total grams per year per state and dispenser/retailer was accessed through the publicly available ARCOS Report 5. ARCOS showed very high correspondence with state Prescription Drug Monitoring Programs and has been widely used in prior pharmacoepidemiology investigations (7, 8, 9, 10, 11). This study was approved by the Institutional Review Boards of Geisinger and the University of New England.

### Analysis

The programs GraphPad Prism, Microsoft Excel, and JMP were used to graph and analyze the data. As the weight (g) distributed directly to practitioners, mid-level practitioners, and teaching institutions was negligible, the state-level analysis was only completed on pharmacies and hospitals. We totaled pharmacy and hospital weights of morphine for each year per state and divided them by the population of the state for a population-adjusted calculation for 2012, which was the peak year of total morphine distributed, and 2020. Population information for 2012 and 2020 was obtained from the US Census Bureau. Then, the percent change between 2012 and 2020 was calculated for each state using their respective population adjusted weights. The same procedure was repeated for percent changes for hospitals alone and pharmacies alone. States with a percent change outside a 95% confidence interval (mean ± 1.96 * Standard Deviation) were considered statistically significant (*p* < .05).

## Results

The peak year for total morphine distribution between 2010 and 2020 was 2012 (24.2 metric tons). This decreased by -55.6% in 2020 (10.7 metric tons, Figure 1).

**Figure 1.**
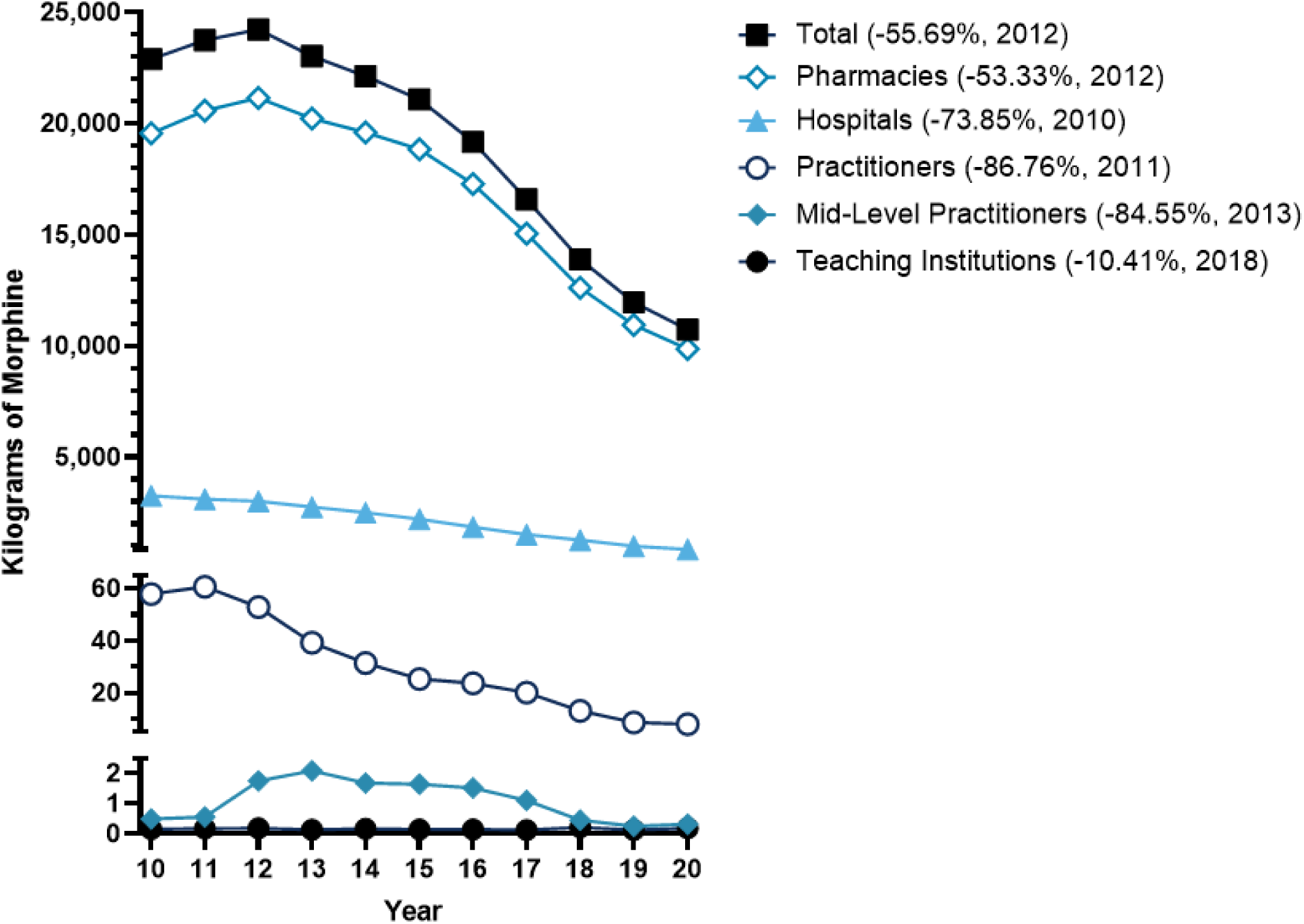
Morphine distribution by business types from 2012 to 2020, as reported by the Drug Enforcement Administration’s Automated Reports and Consolidated Ordering System (ARCOS). The percent change relative to the peak year is shown in parentheses.

Pharmacies accounted for the preponderance of morphine prescribed by any business type in 2012 (87.3%) which increased even further in 2020 (92.0%). Hospitals peak for morphine (2010) was earlier than pharmacies (2012).

Pharmacy and hospital opioid distribution were broken down by state, by the morphine weight distributed per person by each state’s population (Figure 2). Tennessee distributed the highest (180.2 mg/person) and Texas the lowest (39.4 mg/person) in 2012 – a 4.5-fold difference. In 2020, Tennessee remained the state distributing the highest amount of morphine per person (56.36 mg/person) while the District of Columbia was the lowest (14.97 mg/person) – a 3.7-fold difference (Fig S1).

**Figure 2.**
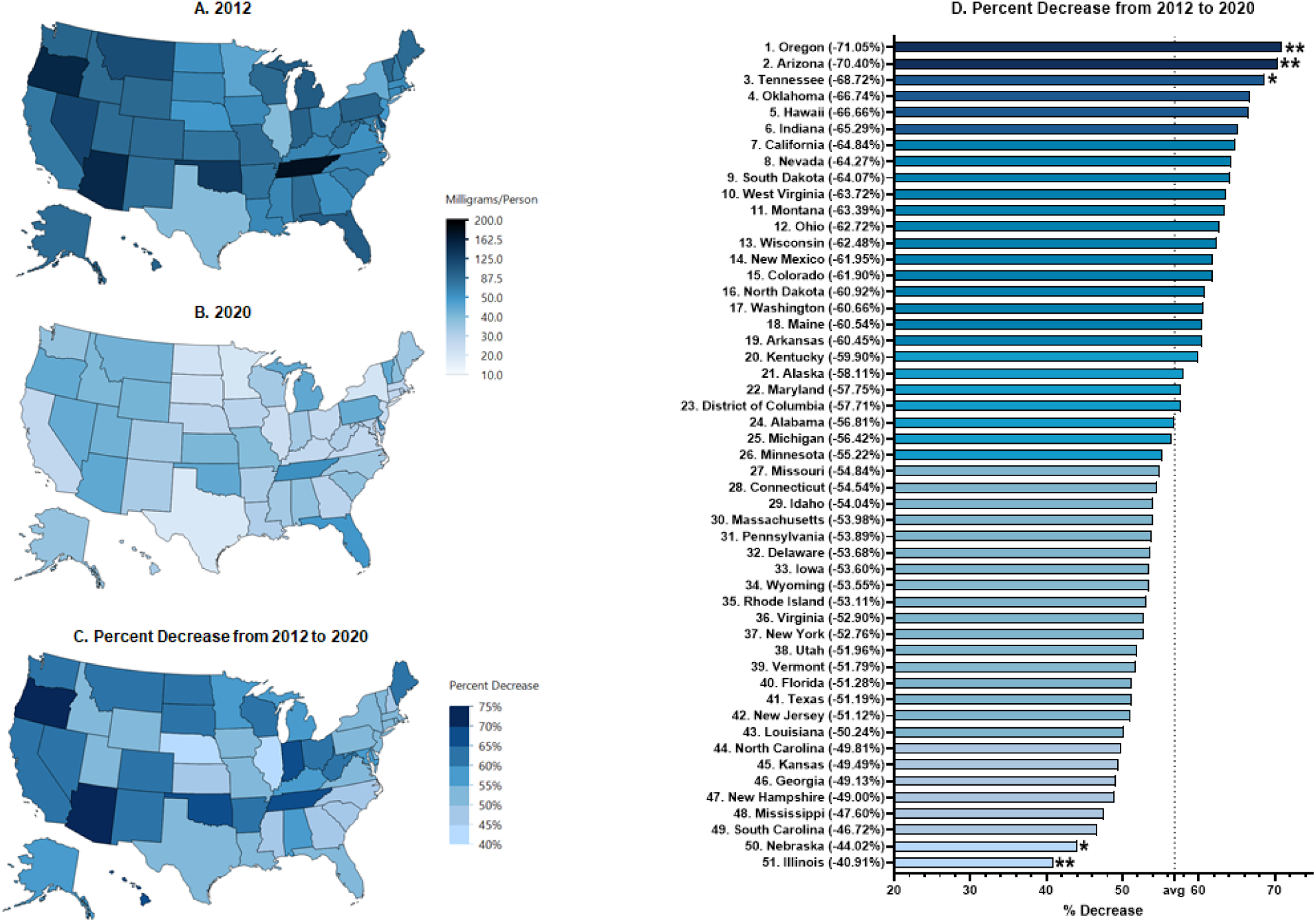
Morphine distribution to pharmacies and hospitals in milligrams per person per state in 2012 **(A)** and 2020 **(B)** as reported by the Drug Enforcement Administration’s Automated Reports and Consolidated Ordering System (ARCOS). Hea map **(C)** and state ranking **(D)** Percent decrease from 2012 to 2020 as reported in **A** and **B**. Average decrease was -56.82%. *State values outside of *1.50 or **1.96 Standard Deviations.

Several changes were notable between 2012 and 2020. First, the combined hospital and pharmacy national distribution declined by -56.4%. Second, in 2012, two of the top three morphine prescribing states, Oregon (154.8 mg/person) and Arizona (153.3 mg/person) saw the largest decreases in distribution between 2012 and 2020 (−71.1% for Oregon and -70.4% for Arizona). Thirdly, Illinois distributed 40.0 mg/person in 2012 and 23.61 mg/person in 2020. This state’s decrease (40.9%) was significantly smaller than the mean reduction (−56.8%). Notably, although pharmacies accounted for more morphine distributed by any business type in 2012, hospital decrease (−72.7%) from 2012-2020 was larger than that of pharmacies (−56.12%, Figures S2 and S3).

## Discussion

This study identified three key findings about the US distribution of morphine over the past decade. First, total morphine distribution from hospitals and pharmacies substantially decreased (−56.4%) since peaking in 2012. Second, the size of this reduction varied across the 50 states and Washington D.C., with an approximately two-fold difference between the largest (OR = -71.1%) and smallest (IL = -40.9%) decreases. Third, despite the overall reduction, a four-fold difference in population corrected use of morphine was observed between states which persisted over the decade.

The overall reduction in morphine distribution nationally coincides with more aggressive and comprehensive policies and initiatives prioritizing the opioid crisis as a healthcare issue, including the US Department of Health and Human Services’ $1 billion fund allocated to states to be used for addiction prevention, treatment, and recovery services, data collection, pain management, overdose reversing drugs such as naloxone, and research (13). Usage of these funds has largely lied within individual state legislation (13).

The US DEA decreased quotas after the passage of the SUPPORT Act, which called on the DEA to quantify diversion of prescription opioids and “make appropriate quota reductions” (14). In 2012, the DEA quota for morphine production (for sale) was 48,200 kilograms; this was decreased by 42.4% to 27,784 kilograms in 2020 (15, 16). Interestingly, this reduction in the overall production quotas of opioids is concurrent with the rise in the production of marijuana (14,17). Indeed, another factor contributing to the decline in morphine prescriptions may be the nationwide shortage of parenteral opioids (notably morphine, hydromorphone, and fentanyl) that has resulted in a transition to alternative opioids such as nalbuphine and buprenorphine (18).

The variability among different states with regard to their success in reducing morphine prescriptions may be attributable to differences in state policies in handling the opioid crisis. In 2020, pharmacies and hospitals in Texas distributed 19.21 mg/person of morphine, the smallest population-adjusted amount out of all fifty states excluding Washington D.C. The decline in Texas’ prescription rates of morphine and all other opioids in general have been attributed to the implementation of several state policies, including the “pill mill” law passed in 2010 that led to significant decreases in monthly morphine prescription volume (19). Oregon is another state in which morphine prescriptions have been reduced through policy efforts. The Oregon Health Authority launched The Opioid Initiative in 2015, which works to increase access to nonopioid pain treatment, support medication-assisted treatment and naloxone access for those using opioids, decrease opioid prescribing, and use data to inform policies and interventions (20). From 2015 to 2017, the number of Oregonians on 90 or more Morphine Equivalent Doses (MEDs) decreased by 37%, from 11.1 per 1000 residents quarterly to 7.0 per 1000 residents quarterly. Prescription opioid overdose deaths decreased 20% from 4.5 per 100 000 in 2015 to 3.6 per 100 000 in 2016 (20). Policies limiting opioid prescriptions (regulations for prescribing to “high-risk” Medicaid payers, required urine drugs tests and documentation of justification for high-dose opioid prescriptions) may account for the success of Oregon’s Opioid Initiative, especially as 4.1% of all prescribers were responsible for 60% of controlled substance prescriptions (21).

It did not escape notice that despite pronounced declines in morphine distribution during the past decade, state level variation was both considerable and consistent over time. For comparison, the state-level variation in 2016 for the MME total of eight opioids used for pain was 3.6 fold between Tennessee (932 MME/person) and Washington D.C. (256 MME/person) (7). The difference for individual opioids was 4.3 fold for fentanyl to hospitals, 6.2 for fentanyl to pharmacies (9), but 18-fold for meperidine (8) and almost 20-fold different for buprenorphine (11). We do not believe that there are four-fold biological nociceptive differences between the residents of Tennessee relative to those in an adjacent state (e.g., Kentucky) that receive much less morphine. Future research should continue to explore the provider (22) or patient attitudes (23), insurance company (24) or pharmacy policies (25) that would account for this sizable variation. This might aid in the identification of practices that are incongruent with evidence-based medicine and warrant improved efforts to balance the benefits of morphine (e.g. for acute pain, chronic cancer pain, and palliative care) with the appreciable risks for this Schedule II substance.

A strength of this study was that it consolidated both hospital and pharmacy morphine distributions. This controls for potential inconsistencies in defining “hospital” vs. “pharmacy” prescribed opioids, as postoperative morphine prescriptions that are written in a hospital but filled in an outside pharmacy are considered “pharmacy-distributed” by ARCOS. The ARCOS data is comprehensive and includes public (Veteran’s Affairs) and private health care facilities. A potential limitation of this study is that the drug distribution amounts are listed in weight (grams) rather than number of prescriptions. However, ARCOS showed a very high correlation (r = 0.985) for oxycodone with a state prescription drug monitoring program (12). Also notable was that during the COVID-19 pandemic, prohibitions against sending controlling substances across state lines were relaxed (26). As the state level differences in 2019 (4.7 fold) and 2020 (3.8 fold) were similar, we believe any impact of telemedicine for this opioid was modest. Finally, this data was limited to the US. The United Nation’s International Narcotic Control Board has reported that there are tremendous disparities between countries in prescription opioid use in North America, Western Europe, Australia, relative to all other countries (2).

Future directions for this line of research can include further investigation of the differences that may result in state-by-state variation in the availability of morphine (including brand and generic manufacturers for both oral and IV morphine) and reliance on opioid adjuncts in each state. Characterization of the primary patient populations that are prescribed morphine in each state, as rural residents and Medicaid enrollees are associated with higher rates of morphine prescriptions and adverse effects of morphine usage (29). Although the evidence for any measurable impact of state-level opioid prescribing laws varies between modest (27) and negligible (28), future research should examine any impact of the *2022 CDC Clinical Practice Guideline for Prescribing Opioids* (30).

In conclusion, this study was conducted to observe the national and state-by-state changes in morphine prescriptions by pharmacies and hospitals in the US from 2012, the peak year for opioid distribution, to 2020. Since 2012, morphine prescriptions have decreased in every state, although states with higher morphine distribution in 2012 were observed to have more pronounced decreases in morphine distribution over the 7-year period.

## Supporting information

DEA ARCOS raw morphine data

## Data Availability

Raw data is available at: https://www.deadiversion.usdoj.gov/arcos/retail_drug_summary/index.html

## Acknowledgments

Thanks to Iris Johnston for technical support. BJP is supported by the Health Resources Services Administration (D34HP31025).

---

**Figure S1.**
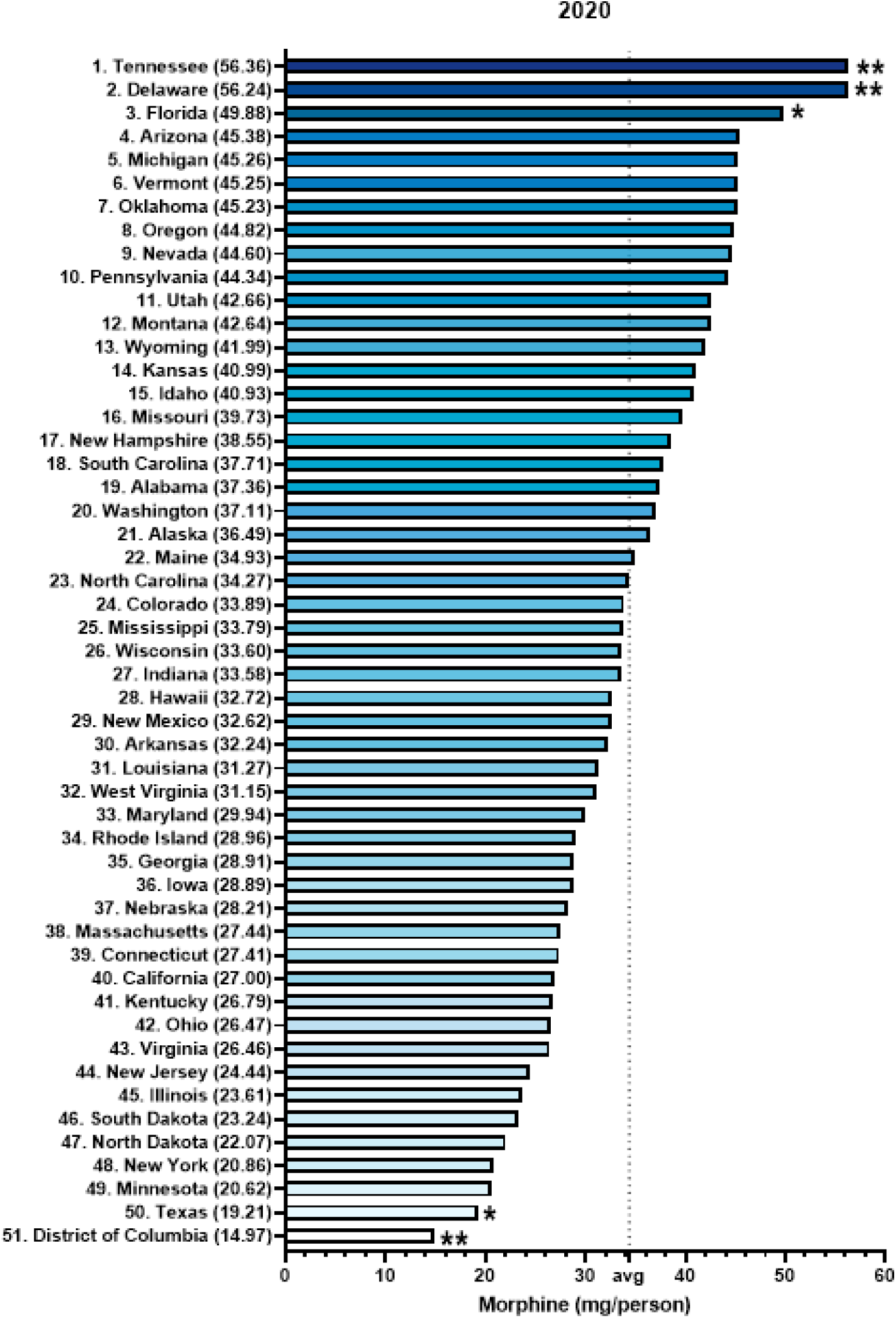
Total pharmacy and hospital distributed morphine in milligrams per person per state in 2020 as reported by the US Drug Enforcement Administration’s Automated Reports and Consolidated Ordering System (ARCOS). States outside *1.5 or **1.96 Standard Deviations (*p* < .05).

**Figure S2.**
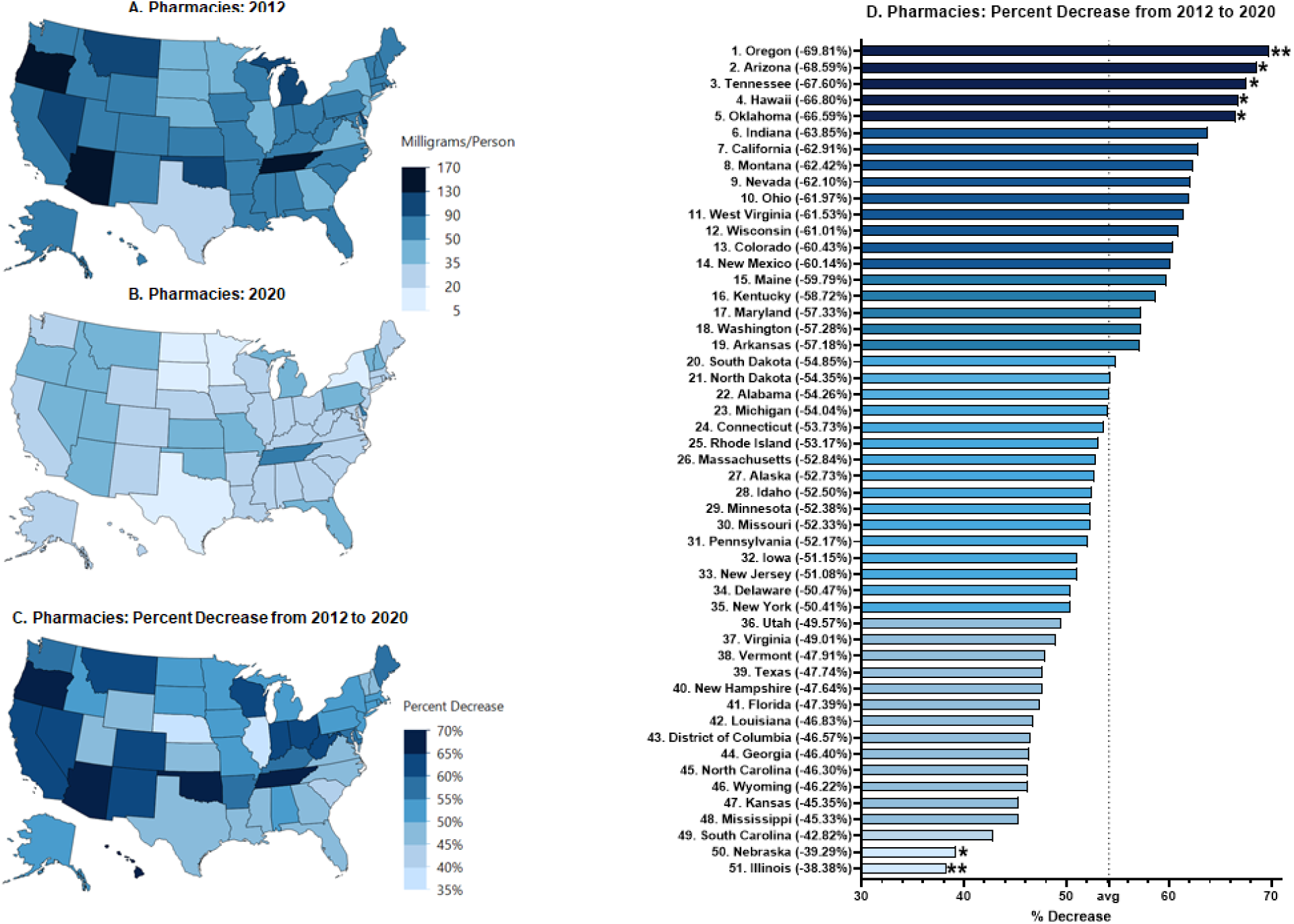
Pharmacy-distributed morphine in milligrams per person per state in 2012 **(A)** and 2020 **(B)** as reported by the Drug Enforcement Administration’s Automated Reports and Consolidated Ordering System (ARCOS). Heatmap **(C)** and state ranking **(D)** of percent decrease from 2012 to 2020. The mean decrease was -54.14%. State values outside *1.50 or **1.96 Standard Deviations (*p* < .05).

**Figure S3.**
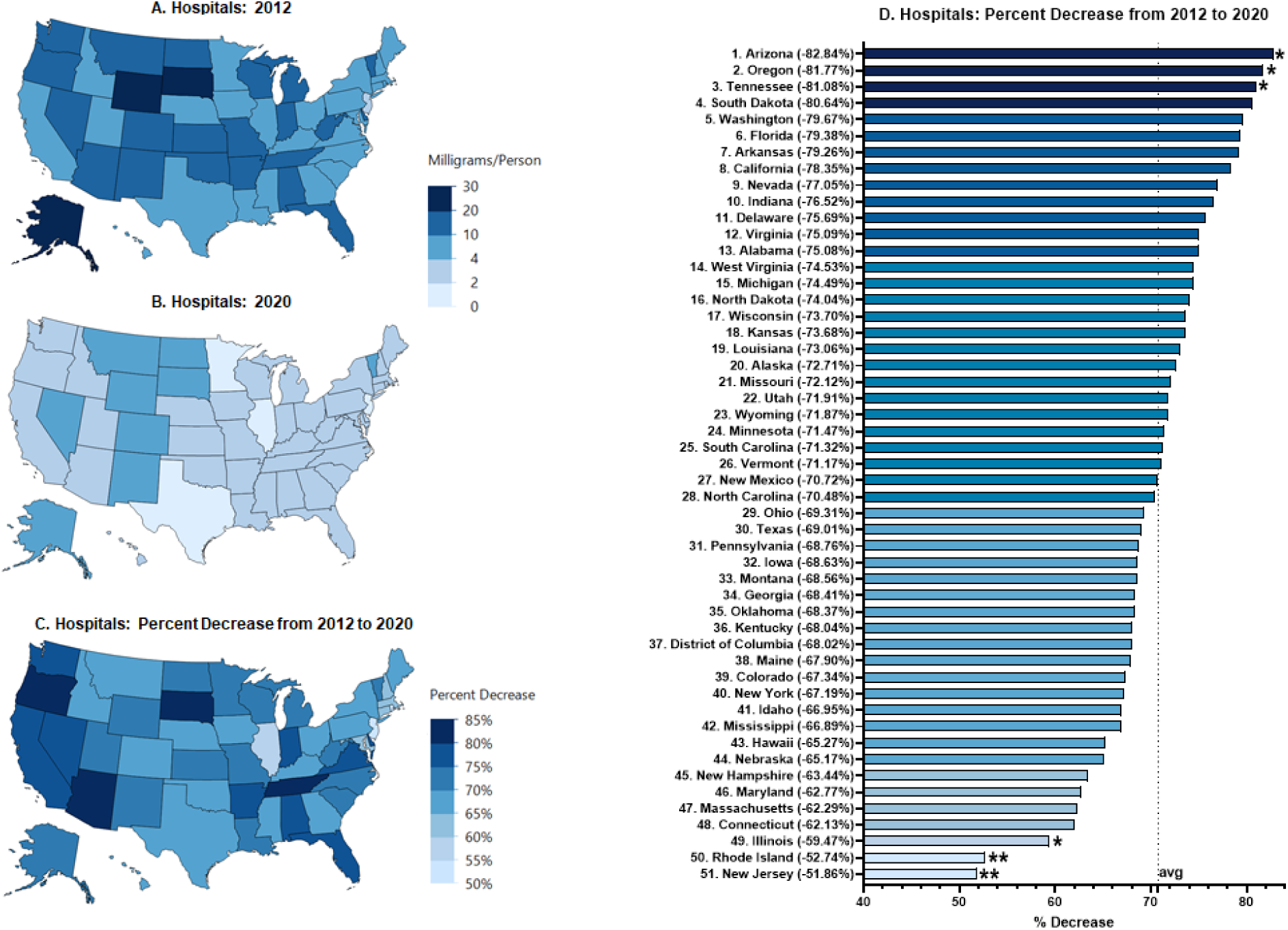
Hospital-distributed morphine in milligrams per person per state in 2012 **(A)** and 2020 **(B)** as reported by the Drug Enforcement Administration’s Automated Reports and Consolidated Ordering System (ARCOS). Heatmap **(C)** and ranking **(D)** of percent decrease from 2012 to 2020. The mean decrease was -70.75%. State values outside *1.50 or **1.96 Standard Deviations (*p* < .05)

## Notes

**Conflict of interest:** BJP was (2019-2021) part of an osteoarthritis research team supported by Pfizer and Eli Lilly. The other authors have no disclosures.

### Competing Interest Statement

BJP was (2019-2021) part of an osteoarthritis research team supported by Pfizer and Eli Lilly. The other authors have no disclosures.

### Funding Statement

This study did not receive any external funding. Publication costs may be paid for by Geisinger Commonwealth School of Medicine. BJP is supported by the Health Resources Services Administration (D34HP31025).

### Author Declarations

The study used ONLY openly available data that were originally located: https://www.deadiversion.usdoj.gov/arcos/retail_drug_summary/index.html

